# Effectiveness of XBB.1.5 Vaccines Against Omicron Subvariants

**DOI:** 10.1101/2024.07.01.24309806

**Authors:** Dan-Yu Lin, Yi Du, Yangjianchen Xu, Sai Paritala, Matthew Donahue, Patrick Maloney

**Affiliations:** Department of Biostatistics, Gillings School of Global Public Health, University of North Carolina at Chapel Hill, Chapel Hill, NC, USA; Division of Public Health, Nebraska Department of Health and Human Services, Lincoln, NE, USA; Department of Epidemiology, College of Public Health, University of Nebraska Medical Center, Omaha, NE, USA

**Author notes:** Correspondence to: Professor Dan-Yu Lin, Department of Biostatistics, University of North Carolina at Chapel Hill, 3101E McGavran-Greenberg Hall, Chapel Hill, NC 27599-7420, USA, Professor Patrick Maloney, Department of Epidemiology, College of Public Health, University of Nebraska Medical Center, Omaha, NE 68198-4395, USA.

## Abstract

**Background:** The updated Moderna, Pfizer-BioNTech, and Novavax COVID-19 vaccines containing the SARS-CoV-2 omicron XBB.1.5 strain have replaced their predecessors in the United States and in other countries since the fall of 2023. The clinical impact of these vaccines on currently circulating variants was unknown.

**Aims:** We aimed to assess the effectiveness of the updated XBB.1.5 vaccines against currently circulating omicron subvariants.

**Methods:** We examined data on the administration of XBB.1.5 vaccines and the incidence of COVID-19 between September 11 and November 27, 2023 for approximately 2 million persons by linking records from the Nebraska Electronic Disease Surveillance System and the Nebraska State Immunization Information System. We used Cox regression to estimate the effects of XBB.1.5 vaccines on the risk of COVID-19, as a function of time elapsed since vaccination, while adjusting for demographic factors, previous infection history, and previous vaccination history.

**Results:** The effectiveness (i.e., proportionate reduction of risk) for XBB.1.5 vaccines against SARS-CoV-2 infection was 63.0% (95% confidence interval [CI], 48.6 to 73.4) 4 weeks after vaccination and 67.1% (95% CI, 49.9 to 78.4) 6 weeks after vaccination; vaccine effectiveness started to decline after 6 weeks. Vaccine effectiveness was broadly similar across subgroups defined by age, sex, race and ethnicity, socioeconomic status, and previous immunity status.

**Conclusion:** XBB.1.5 vaccines were effective against currently circulating variants, regardless of age, sex, race and ethnicity, socioeconomic status, or previous immunity status. These findings can be used to develop effective prevention strategies against COVID-19.

## Introduction

Coronavirus disease 2019 (COVID-19) has resulted in 7 million deaths in the world. Vaccination has played a critically important role in combating this pandemic. Vaccine effectiveness wanes over time and tends to be lower against new variants. Knowledge about vaccine effectiveness is useful in developing effective prevention strategies against COVID-19.

On September 11, 2023, the U.S. Food and Drug Administration (FDA) authorized the updated Moderna and Pfizer-BioNTech mRNA COVID-19 vaccines that contain a monovalent component corresponding to the SARS-CoV-2 omicron subvariant XBB.1.5 for all doses administered to individuals 6 months of age and older.^1^ On October 3, 2023, the FDA authorized the updated Novavax COVID-19 Vaccine, Adjuvanted containing the spike protein from the XBB.1.5 subvariant for use in individuals 12 years of age and older.^2^ The FDA no longer authorized the Moderna and Pfizer-BioNTech bivalent vaccines and the original Novavax COVID-19 Vaccine, Adjuvanted for use. The FDA’s decisions were based on the evaluation of manufacturing data and non-clinical immune response data on the updated formulations with the XBB.1.5 component.^1–2^ As of November 2023, there were no published data on the clinical efficacy of these updated vaccines.

We examined individual-level data on the administration of the three XBB.1.5 vaccines and the incidence of COVID-19 in the entire state of Nebraska, with a population of approximately 2 million residents, from September 11, 2023 through November 27, 2023. During this period, the dominant circulating variants in the region changed from EG.5, XBB.2.3, and XBB.1.16 to HV.1 and JN.1, and the proportion of XBB.1.5 declined from 10% to 1% (https://covid.cdc.gov/covid-data-tracker/#variant-proportions). We evaluated the effects of the three vaccines on the risks of SARS-CoV-2 infection and death in the entire population and in subgroups defined by age, sex, race and ethnicity, socioeconomic status, and previous immunity status.

## Methods

### Data Sources

#### Nebraska Electronic Disease Surveillance System

The Nebraska Electronic Disease Surveillance System (NEDSS) is a statewide, web-based infectious disease surveillance and case management system used by the Nebraska Department of Health and Human Services and 19 local health departments covering 93 counties. The NEDSS receives COVID-19 test results, including polymerase-chain-reaction and antigen tests, from facilities participating in electronic laboratory reporting across Nebraska. During the study period, individuals who tested for COVID-19 usually had symptoms consistent with COVID-19. Potential COVID-19 deaths were identified through vital records review and verified through investigations at local health departments using an independent review of death certificates, lab reports, and consultation with physicians, coroners, and/or patient relatives.

#### Nebraska State Immunization Information System

The Nebraska State Immunization Information System (NESIIS) is a statewide, web-based immunization information system that secures COVID-19 vaccine data from public clinics, private provider offices, local health departments, and hospitals. All Nebraska residents 6 months of age or older were eligible to receive the updated XBB.1.5 vaccines. The NESIIS keeps accurate, up-to-date COVID-19 vaccination records, including types of vaccine (monovalent, bivalent, updated XBB.1.5), dates of vaccine administration, vaccine manufacturers (Moderna, Pfizer-BioNTech, Novavax), number of doses, and vaccine recipient information.

### Data Linkage

We probabilistically matched laboratory and vaccination records by first name, last name, middle name, date of birth, sex, and residential zip code. We used Match*Pro v2.3 software to link the data. The linkage was based on the Fellegi and Sunter model, under which a probabilistic score was used to determine the quality of the match. Records that demonstrated a high total match score, typically above 29.0, were considered to belong to the same individual. Case investigations were linked directly using a unique identifier. The linked records were also reviewed manually.

### Analysis Dataset

The analysis dataset was prepared by merging data on COVID-19 laboratory testing results, COVID-19 vaccination records, vital records, and COVID-19 case investigation records with death information from November 27, 2022 to November 27, 2023, for Nebraska residents with records of vaccination or COVID-19 testing. Demographic data were obtained from the NEDSS and NESIIS and included in the linked dataset. Missing values in demographic variables were filled using a hot-deck single imputation method.

We used the Nebraska Department of Health and Human Services (DHHS) population master database to create dummy records on age, sex, race, ethnicity, and county of residence for persons who were not in the NESIIS or NEDSS database (i.e., no records of COVID-19 vaccination or positive COVID-19 diagnosis). This population master database was carefully integrated from various sources, including COVID-19 laboratory test and case investigation records from the NEDSS, COVID-19 vaccination records from the NESIIS, driver license database from the Nebraska Department of Motor Vehicles, birth certificates and death certificates from the Nebraska Vital Records System, cancer database from the Nebraska Cancer Registry, and hospital discharge records from the Nebraska Hospital Association.

### Statistical Analysis

We fit a Cox regression model in which the hazard ratio of SARS-CoV-2 infection for the updated vaccine depends on the time elapsed since injection.^3–5^ We used a piecewise linear function for the log hazard ratio with change points at 2, 4, and 6 weeks (i.e., 14, 28, and 42 days) after vaccination to ascertain the ramping-up and waning effects of the updated vaccine. In addition, we let the hazard ratio be piecewise constant with change points at 2 and 4 weeks to estimate average vaccine effects over successive time periods (Supplementary Methods).

To reduce confounding bias due to time trends in disease incidence, we measured the time to SARS-CoV-2 infection for each person from a common origin, namely September 11, 2023, such that the risks of disease for vaccinated and unvaccinated persons are compared on the same calendar date. To further reduce confounding bias, we included the following baseline characteristics as covariates: time since previous vaccination, which was set to 9 months if it was greater than 9 months or if the person was not previously vaccinated; time since previous infection, which was set to 9 months if it was greater than 9 months or if the person was not previously infected; and the demographic factors of sex, age group, race and ethnicity, and socioeconomic status.

We created a socioeconomic index by zip code according to median income, fraction of vacant housing, fraction of poverty, fraction of persons without health insurance, fraction of persons with a high school degree or higher, and fraction of persons with assisted income, with a higher index indicating worse socioeconomic standing.^6^ We then dichotomized this index at the sample median to create the “low” versus “high” categories. In addition, we created a race and ethnicity variable by contrasting non-Hispanic white with all others.

We defined vaccine effectiveness as 100x(1–HR)%, where HR is the hazard ratio under the aforementioned Cox regression model. In addition, we defined vaccine effectiveness with internal control as 100x(1–HR3/HR1), where HR1 and HR3 are the hazard ratios in weeks 1–2 and weeks 5–10 after vaccination, respectively. This quantity removes the confounding bias due to non-random administration of updated vaccines and provides a lower bound for the causal effects of updated vaccines (Supplementary Methods). Maximum partial likelihood was used to estimate the hazard ratio and the two measures of vaccine effectiveness and construct 95% confidence intervals (CIs).

We estimated vaccine effectiveness for the three updated vaccines combined and also for the Moderna and Pfizer-BioNTech vaccines separately. In addition, we estimated vaccine effectiveness for the subgroups defined by age, sex, race and ethnicity, socioeconomic status, and previous immunity status. The last variable compared persons who had been vaccinated or infected (or both) within the past 9 months with those who had not been vaccinated or infected within the past 9 months.

Because the number of deaths was small, we estimated an overall vaccine effectiveness against death by assuming a time-constant hazard ratio for the XBB.1.5 vaccines. We adjusted only for demographic variables, because none of the previously infected persons died and only one of the previously vaccinated persons died.

## Results

### Study Population

Table 1 summarizes the demographic characteristics of the Nebraskan population, together with vaccine uptake and clinical outcomes in the state from September 11, 2023 to November 27, 2023. In addition, Figure 1 shows the variant proportions over time, together with the time trends for vaccine uptake and SARS-CoV-2 infection.

**Figure 1.**
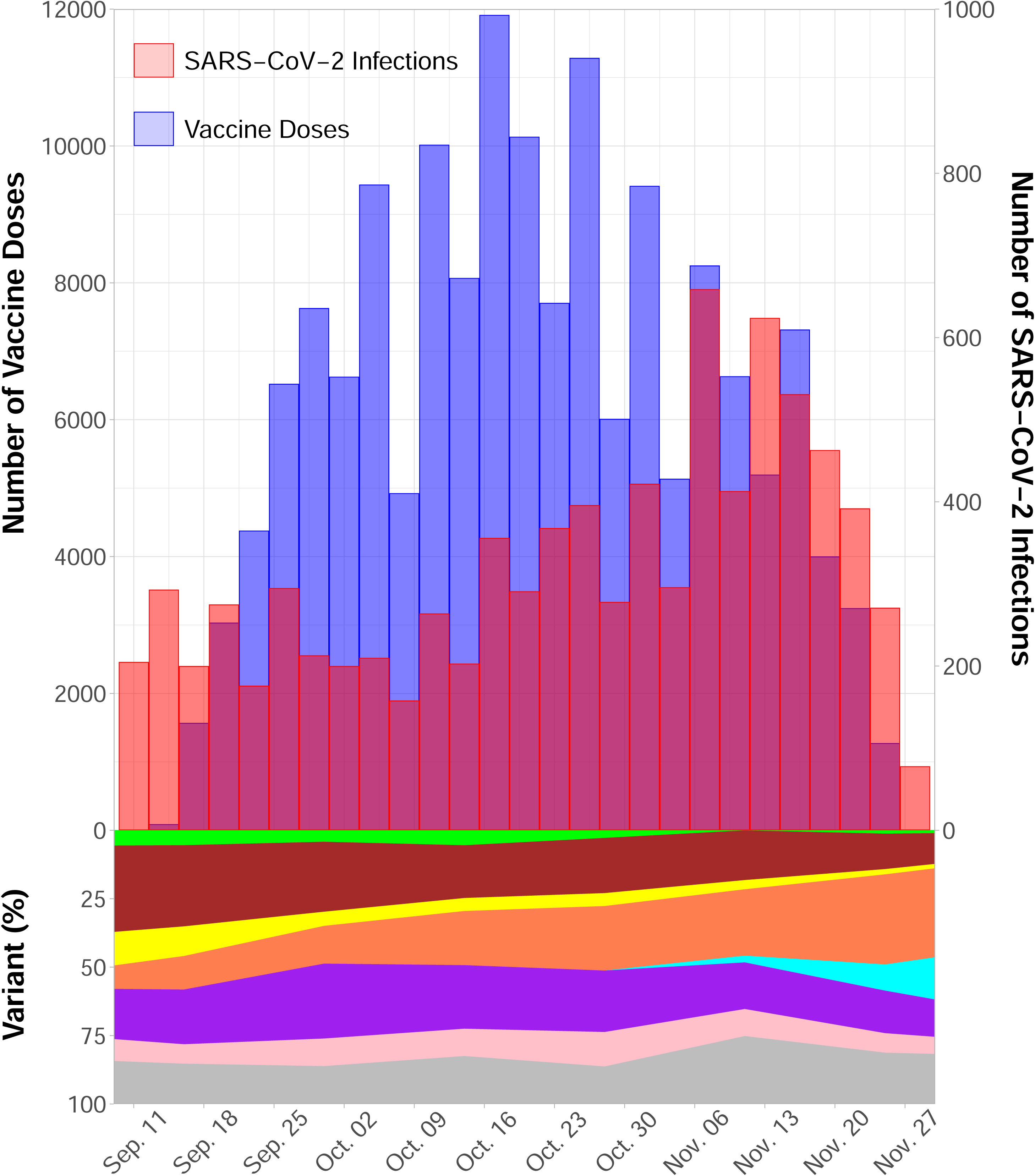
Number of persons receiving XBB.1.5 vaccines, number of SARS-CoV-2 infections, and proportions of circulating variants from September 11 to November 27, 2023 in the state of Nebraska. Omicron subvariants XBB.1.5 and its extensions, XBB.1.16 and its extensions, XBB.2.3 and its extensions, HV.1, JN.1, EG.5, FL.1.5.1, and all other subvariants are indicated by green, brown, yellow, coral, cyan, purple, pink, and grey, respectively. Variant proportions were based on the viral genomic surveillance data for Region 7 (Iowa, Kansas, Missouri, and Nebraska) from the U.S. Centers for Disease Control and Prevention, available at https://covid.cdc.gov/covid-data-tracker/#variant-proportions.

**Table 1.** Demographic Characteristics of the Nebraska Population According to Vaccine Uptake and Clinical Outcomes from September 11, 2023, to November 27, 2023.

| Characteristic | Overall |  | Vaccine Uptake |  |  | Clinical Outcomes |  |
| --- | --- | --- | --- | --- | --- | --- | --- |
|  |  |  | Moderna | Pfizer-BioNTech | Novavax | Infection | Death<sup>a</sup> |
|  | No. (column %) |  |  | No. (row %) |  | No. (row %) |  |
| All persons | 2,090,367 (100) |  | 61,435 (2.94) | 98,034 (4.69) | 370 (0.018) | 8,530 (0.41) | 39 (0.0019) |
| Age |  |  |  |  |  |  |  |
| 0-11 | 190,403 (9) |  | 2,782 (1.46) | 1,401 (0.74) | 0 (0) | 705 (0.37) | 0 (0) |
| 12-64 | 1,417,409 (68) |  | 23,569 (1.66) | 46,606 (3.29) | 258 (0.018) | 4,586 (0.32) | 2 (0.0001) |
| ≥65 | 482,555 (23) |  | 35,084 (7.27) | 50,027 (10.37) | 112 (0.023) | 3,239 (0.67) | 37 (0.0077) |
| Sex |  |  |  |  |  |  |  |
| Male | 996,605 (48) |  | 26,829 (2.69) | 42,126 (4.23) | 153 (0.015) | 3,248 (0.33) | 16 (0.0016) |
| Female | 1,093,762 (52) |  | 34,606 (3.16) | 55,908 (5.11) | 217 (0.020) | 5,282 (0.48) | 23 (0.0021) |
| Race and ethnicity |  |  |  |  |  |  |  |
| Non-Hispanic white | 1,633,705 (78) |  | 54,990 (3.37) | 86,673 (5.31) | 347 (0.021) | 6,923 (0.42) | 37 (0.0023) |
| All others | 456,662 (22) |  | 6,445 (1.41) | 11,361 (2.49) | 23 (0.005) | 1,607 (0.35) | 2 (0.0004) |
| Socioeconomic status |  |  |  |  |  |  |  |
| Low | 1,176,458 (56) |  | 32,414 (2.76) | 53,697 (4.56) | 145 (0.012) | 4,538 (0.39) | 17 (0.0015) |
| High | 913,909 (44) |  | 29,021 (3.18) | 44,337 (4.85) | 225 (0.025) | 3,992 (0.44) | 22 (0.0024) |
| Previous vaccination |  |  |  |  |  |  |  |
| <9 months | 92,400 (4) |  | 8,222 (8.90) | 13,706 (14.83) | 38 (0.041) | 534 (0.58) | 1 (0.0011) |
| ≥9 months | 1,997,967 (96) |  | 53,213 (2.66) | 84,328 (4.22) | 332 (0.017) | 7,996 (0.40) | 38 (0.0019) |
| Previous infection |  |  |  |  |  |  |  |
| <9 months | 27,158 (1) |  | 1,148 (4.23) | 1,817 (6.69) | 4 (0.015) | 235 (0.87) | 0 (0) |
| ≥9 months | 2,063,209 (99) |  | 60,287 (2.92) | 96,217 (4.66) | 366 (0.018) | 8,295 (0.4) | 39 (0.0019) |

a. Identified through vital records.

Nebraska has a diverse population that reflects the age, sex, and income distributions of the United States. However, the percentage of Black or African American and the percentage of persons who identify as Hispanic, Asian, or Pacific Islander in Nebraska are lower than the national averages.^7^

Vaccination rates were higher among older adults, females, and non-Hispanic white persons. The Pfizer-BioNTech vaccine was the most commonly administered, whereas very few people received the Novavax vaccine. Vaccine uptake peaked in the month of October.

From September 11, 2023 to November 27, 2023, the dominant circulating variants in the region changed from EG.5, XBB.2.3, and XBB.1.16 to HV.1 and JN.1. The proportion of the XBB.1.5 subvariant declined from 10% to 1%.

A total of 8,530 SARS-CoV-2 infections were identified. The linkage to vital records indicated that 39 of those infections resulted in deaths, 12 of which had been verified by case investigations as of November 27, 2023. (Case investigations may take several months to complete, leading to delays in confirmation of deaths identified through vital statistics.) The rates of SARS-CoV-2 infection and death were the highest among persons 65 years of age and older.

### Vaccine Effectiveness Against SARS-CoV-2 Infection

Estimates of vaccine effectiveness in reducing the risk of SARS-CoV-2 infection over time are shown in Figure 2 and Figures S1. For all persons, vaccine effectiveness was 63.0% (95% CI, 48.6 to 73.4), 67.1% (95% CI, 49.9 to 78.4), and 47.1% (95% CI, 20.4 to 64.8) 4 weeks, 6 weeks, and 8 weeks after vaccination, respectively (Fig. 2; Table S1). For the Pfizer-BioNTech vaccine, effectiveness was 75.9% (95% CI, 60.1 to 85.4), 68.3% (95% CI, 44.6 to 81.8), and 56.1% (95% CI, 22.8 to 75.1) 4 weeks, 6 weeks, and 8 weeks after vaccination, respectively; for the Moderna vaccine, effectiveness was 42.1% (95% CI, 10.6 to 62.6), 66.0% (95% CI, 34.6 to 82.3), and 31.2% (95% CI, -23.5 to 61.7) 4 weeks, 6 weeks, and 8 weeks after vaccination, respectively (Fig. S1 A; Table S2). We were unable to obtain a stable estimate for the effectiveness of the Novavax vaccine because very few people received this updated vaccine and because there was only one SARS-CoV-2 infection after injection.

**Figure 2.**
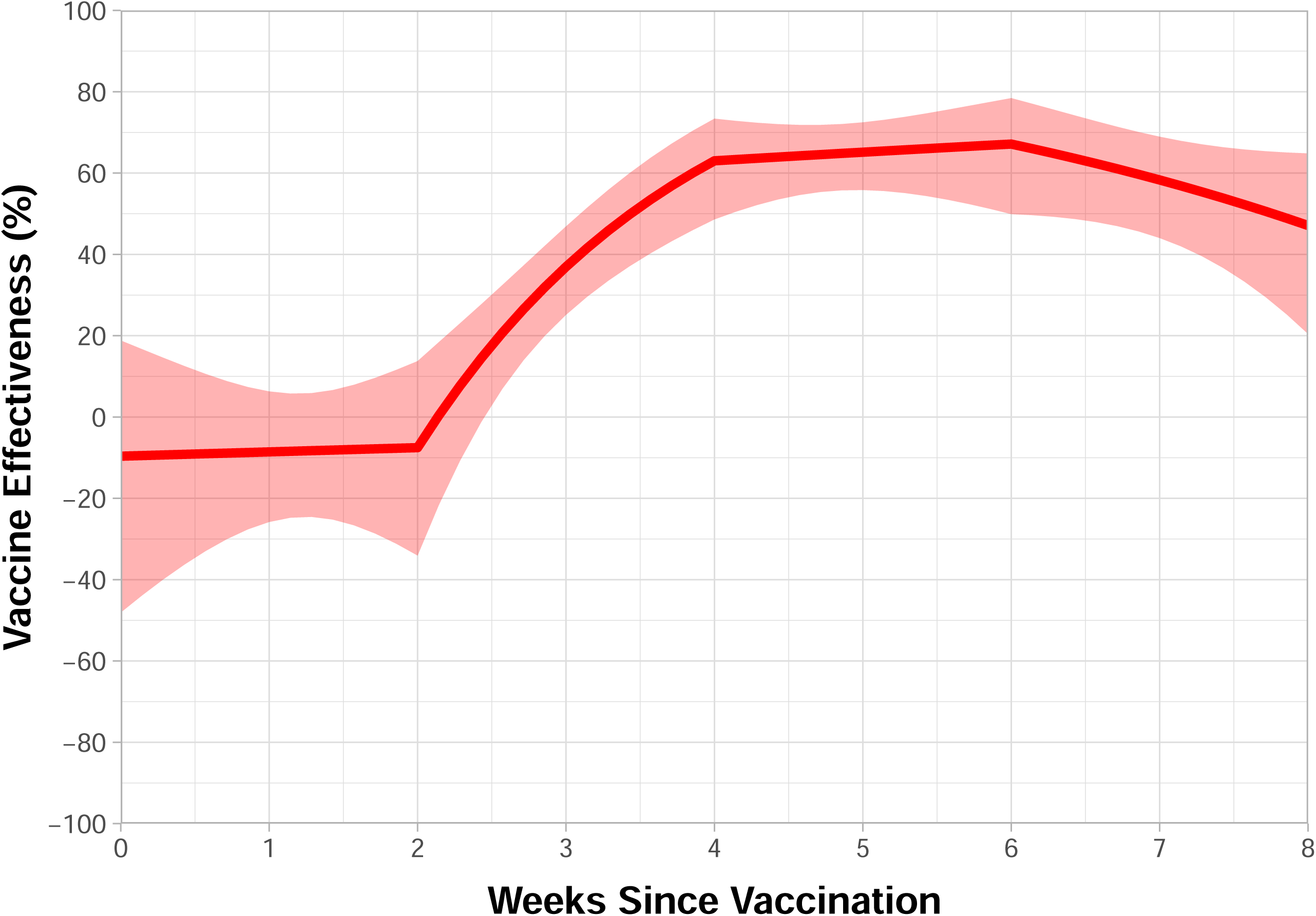
Effectiveness of XBB.1.5 vaccines in reducing the risk of SARS-CoV-2 infection as a function of time since vaccination. Estimates are shown by solid curves, and 95% confidence intervals are indicated by shaded bands.

For persons 12–64 years of age, vaccine effectiveness was 48.7% (95% CI, 10.3 to 70.7), 39.1% (95% CI, -16.2 to 68.1), and 32.0% (95% CI, -40.9 to 67.2) 4 weeks, 6 weeks, and 8 weeks after vaccination, respectively; for persons 65 years of age and older, vaccine effectiveness was 68.5% (95% CI, 52.5 to 79.0), 76.4% (95% CI, 58.9 to 86.5), and 52.5% (95% CI, 22.1 to 71.0) 4 weeks, 6 weeks, and 8 weeks after vaccination, respectively (Fig. S1 B; Table S2). We were unable to obtain a stable estimate of the vaccine effectiveness for children under 12 years of age because there were only two SARS-CoV-2 infections after vaccine receipt in that age group.

Vaccine effectiveness was broadly similar between males and females, between non-Hispanic whites and other race and ethnicity groups, between persons of low and high socioeconomic standing, and between persons who were vaccinated or infected within the past 9 months and those who were not vaccinated or infected with the past 9 months (Fig. S1 C–F; Tables S3–S4). Due to smaller sample sizes, the confidence intervals for subgroups are wider than those of all persons.

Estimates of average vaccine effectiveness over successive time periods and estimates of vaccine effectiveness with internal control are shown in Table 2. Because very few people had been vaccinated more than 10 weeks ago, the estimates of average vaccine effectiveness after the changepoint of 4 weeks pertain to weeks 5–10 after vaccination. For all persons, average vaccine effectiveness in weeks 5–10 was estimated at 60.0% (95% CI, 50.1 to 67.9), and the corresponding vaccine effectiveness with internal control was estimated at 65.1% (95% CI, 54.4 to 73.2). The reason that the latter estimate was slightly higher than the former was because vaccinated persons had slightly greater risk of SARS-CoV-2 infection over the first two weeks after vaccination than unvaccinated persons, which implies that persons at higher risk for SARS-CoV-2 infection were slightly more likely to be vaccinated.

**Table 2.** Estimates and 95% Confidence Intervals for the Effectiveness of One XBB.1.5 Vaccine Dose Against SARS-CoV-2 Infection.

|  | **Weeks 1–2** | **Weeks 3–4** | **Weeks 5–10** | **Weeks 5–10 vs 1–2<sup>a</sup>** |
| --- | --- | --- | --- | --- |
| All Persons | -14.4% (-33.3 to 1.7) | 38.2% (24.4 to 49.4) | 60.0% (50.1 to 67.9) | 65.1% (54.4 to 73.2) |
| Manufacturer |  |  |  |  |
| Pfizer-BioNTech | 7.3% (-15.2 to 25.4) | 56.7% (41.1 to 68.2) | 66.4% (54.6 to 75.2) | 63.8% (47.6 to 75.0) |
| Moderna | -47.1% (-81.5 to -19.3) | 9.4% (-18.0 to 30.4) | 50.1% (31.0 to 63.9) | 66.1% (50.2 to 76.9) |
| Age |  |  |  |  |
| 12–64 | 0.9% (-33.9 to 26.7) | 25.6% (-5.4 to 47.5) | 44.9% (20.4 to 61.8) | 44.4% (10.8 to 65.3) |
| ≥65 | -20.7% (-44.1 to -1.0) | 45.2% (29.6 to 57.4) | 65.5% (54.4 to 73.8) | 71.4% (60.4 to 79.3) |
| Sex |  |  |  |  |
| Male | -16.1% (49.3 to 9.7) | 40.8% (16.8 to 57.8) | 61.6% (44.5 to 73.5) | 66.9% (48.6 to 78.7) |
| Female | -13.7% (-37.7 to 6.1) | 36.5% (18.5 to 50.6) | 59.0% (45.9 to 68.8) | 63.9% (49.7 to 74.1) |
| Race and ethnicity |  |  |  |  |
| Non-Hispanic white | -12.1% (-31.5 to 4.4) | 41.9% (28.0 to 53.1) | 60.4% (50.2 to 68.5) | 64.6% (53.5 to 73.1) |
| All others | -26.0% (-110.2 to 24.4) | 0.9% (-79.9 to 45.4) | 62.1% (8.5 to 84.3) | 69.9% (17.2 to 89.1) |
| Socioeconomic status |  |  |  |  |
| Low | 9.7% (-14.9 to 29.1) | 22.9% (0.3 to 40.4) | 67.6% (54.3 to 77.1) | 64.1% (45.5 to 76.3) |
| High | -38.1% (-68.2 to -13.4) | 53.2% (35.2 to 66.2) | 52.1% (36.0 to 64.1) | 65.3% (51.0 to 75.4) |
| Previous immunity status<sup>b</sup> |  |  |  |  |
| High | 18.7% (-22.8 to 46.2) | 58.9% (26.8 to 76.9) | 41.9% (9.8 to 62.5) | 28.5% (-28.3 to 60.2) |
| Low | -24.0% (-46.1 to -5.3) | 32.2% (16.0 to 45.3) | 63.2% (52.4 to 71.6) | 70.3% (59.8 to 78.1) |

- Vaccine effectiveness in reducing the risk of SARS-CoV-2 infection in weeks 5–10 relative to weeks 1–2. - High versus low is defined as vaccinated or infected within the past 9 months versus not vaccinated or infected within the past 9 months.

### Vaccine Effectiveness Against Death

Vital records showed COVID-19 as a primary or contributing cause of death for 39 persons who acquired SARS-CoV-2 infection during the period under study. Only one of the deaths was an XBB.1.5 vaccine recipient. The vaccine effectiveness against death was estimated at 81.2% (95% CI, -38.0 to 97.4). The death of the XBB.1.5 vaccine recipient occurred within 3 weeks after vaccination. Thus, the vaccine effectiveness after the ramping-up period would be estimated at 100%, although with a very wide confidence interval due to the small number of events.

Only 12 deaths had been verified by case investigations, and none of them occurred after receiving XBB.1.5 vaccines. The corresponding estimate of vaccine effectiveness would be 100%, but the confidence interval would be very wide due to the very small number of events.

## Discussion

Although the updated COVID-19 vaccines contain a monovalent component corresponding to the XBB.1.5 strain, we demonstrated that these vaccines were effective against currently circulating omicron subvariants, which included only a small proportion of XBB.1.5. Vaccine effectiveness was moderately high, regardless of a person’s age, sex, race and ethnicity, socioeconomic status, or previous immunity status. The effectiveness of the XBB.1.5 vaccines was lower than the effectiveness of the original monovalent vaccines against the delta variant but was comparable to the effectiveness of the original monovalent boosters against the delta variant and the effectiveness of bivalent boosters against the omicron variant.^3–5, 8–19^

Because the XBB.1.5 vaccines were approved on September 11, 2023 and did not become widely available until October of 2023, we were only able to estimate their effectiveness for about 10 weeks after vaccination, with considerable uncertainties after 8 weeks. We plan to update our data in 3–6 months in order to evaluate the durability of protection afforded by the XBB.1.5 vaccines.

We focused on the endpoint of SARS-CoV-2 infection. Vaccine effectiveness is expected to be higher against hospitalization and death than against infection.^3–5, 8–11^ Indeed, there were greater uncertainties about the effectiveness of the updated XBB.1.5 vaccines against SARS-CoV-2 infection than against hospitalization and death. Thus, our findings are important, and the effectiveness estimates for SARS-CoV-2 infection reported here provide a lower bound for the effectiveness against hospitalization and death.

We were unable to accurately estimate vaccine effectiveness against death from COVID-19 because the number of deaths was small. We did not have real-time access to hospitalization data, so there was a delay in receiving line-level COVID-19 hospitalization data. As a result, hospitalization data were not included in this analysis. However, we plan to retrieve the hospitalization records for our next analysis, by which time the number of deaths may be high enough for us to accurately estimate the effectiveness of the XBB.1.5 vaccines against death.

At-home test results are not reported to the NEDSS. Thus, our database under-represents SARS-CoV-2 infections. The SAR-CoV-2 infections reported to the NEDSS tend to be symptomatic and represent more severe infections, as compared with other infections. Vaccine effectiveness is expected to be lower against asymptomatic and mild infections than against severe infections.

Because vaccine effectiveness was based on relative risk rather than risk difference, under-reporting of SARS-CoV-2 infections would not bias effectiveness estimates unless the likelihood of testing is related to vaccination status.

Reporting of vaccine doses to the NESIIS became optional after the end of the federal declaration of COVID-19 Public Health Emergency on May 11, 2023. However, vaccine providers who participate in the Vaccines for Children or Bridge Access programs and pharmacists are required to report all doses administered. Indeed, the number of providers who reported vaccination data to the NESIIS after September 11, 2023 was about the same as the number of providers who reported vaccination records to the NESIIS before May 11, 2023. Thus, the vaccination information was rather complete. Under-reporting of vaccination could attenuate the estimates of vaccine effectiveness.

As with other observational studies, our analysis was limited by confounding bias. We adjusted for measured confounders (age, sex, race and ethnicity, socioeconomic status, previous infection history, previous vaccination history). In addition, we measured the time to disease occurrence from September 11, 2023 to compare disease incidence between vaccinated and unvaccinated persons on the same date, thus avoiding confounding due to time trends.

The NEDSS and NESIIS do not collect data on underlying medical conditions. Because the FDA recommended individuals who are immunocompromised to be vaccinated at shorter time intervals than those who are immunocompetent. Indeed, our data suggested that persons at higher risks of COVID-19 were more likely to receive the XBB.1.5 vaccines. Thus, lack of adjustment for underlying medical conditions tend to result in underestimation of vaccine effectiveness.

We compared the instantaneous risk of disease between weeks 5–10 and weeks 1–2 among the vaccinated persons. This strategy eliminated confounding bias and provided a lower bound for the causal effects of the XBB.1.5 vaccines. The fact that this measure of vaccine effectiveness was slightly higher than the standard measure of vaccine effectiveness showed that the XBB.1.5 vaccines were indeed effective against currently circulating variants.

### Conclusions

XBB.1.5 vaccines were effective against currently circulating variants, regardless of age, sex, race and ethnicity, socioeconomic status, or previous immunity status. These findings can be used to develop effective prevention strategies against COVID-19.

## Data Availability

Data are available upon the approval of request to the Nebraska Department of Public Health

## Authors’ Contributions

D. Y. Lin designed the study, supervised the analysis, and wrote the paper; Y. Du prepared the analysis dataset; Y. Xu wrote the analysis code, performed the analysis, and generated the tables and figures; S. Paritala performed the analysis; P. Maloney and M. Donahue provided information about the data, guidance and oversight. All authors read and approved of the paper.

## Supplementary Appendix

Supplement to: Lin DY, Du Y, Xu Y, et al. Effectiveness of XBB.1.5 Vaccines Against Omicron Subvariants.

This appendix has been provided by the authors to give readers additional information about the work.

## Supplementary Methods

Let *S* denote the time when the person receives the updated vaccine, and *T* denote the time when the person acquires COVID-19. Both times are measured in days from the start of the study (i.e., September 11, 2023). In addition, let *X* denote baseline risk factors (i.e., age, sex, race/ethnicity, socioeconomic status, time of previous infection, and time of previous vaccination). We specify that the hazard function of *T* is related to *S* and *X* through the Cox regression model

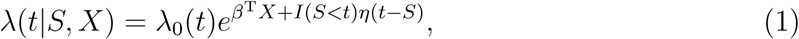

where *I*(*·*) is the indicator function, *λ*_0_(*·*) is an arbitrary baseline hazard function, *β* is a set of log hazard ratios representing the effects of baseline risk factors, and *η*(*·*) is a log hazard ratio function characterizing the time-varying effect of vaccine.^1-3^ Under this model, the level of community transmission may vary over the calendar time, and the effect of vaccine on the risk of COVID-19 depends on the time elapsed since vaccination. Of note, the hazard rate *λ*(*t*) is the probability of disease occurrence at day *t* given that the subject is disease-free before day *t* and represents the risk of disease at day *t* when the disease is rare.

We can approximate the time-varying log hazard ratio *η*(*·*) in model (1) by a continuous piece-wise constant function.^1-3^ The approximation becomes more accurate as the number of change points increases. However, the number of parameters increases with the number of change points. In finite samples, we can obtain stable estimates for only a small number of parameters by choosing a small number of change points. For this study, we set the change points at 2 weeks, 4 weeks, and 6 weeks, which allow us to ascertain the ramping-up and waning effects of the vaccine with sufficient precision and stability. We estimate a continuous function of vaccine effectiveness VE(*t*) = 1 *− e^η^*^(*t*)^.

We also approximate *η*(*·*) by a piecewise constant function with change points at 2 weeks and 4 weeks, which allows us to estimate average vaccine effects over three time intervals after vaccination: weeks 1–2, weeks 3–4, and after 4 weeks. Because very few persons have been vaccinated more than 10 weeks ago, the average vaccine effect after 4 weeks pertain to weeks 5–10.^2^ The average vaccine effectiveness in weeks 1–2, weeks 3–4, and weeks 5–10 are defined by VE_1_ = 1 *−* HR_1_, VE_2_ = 1 *−* HR_2_, and VE_3_ = 1 *−* HR_3_, where HR_1_, HR_2_, and HR_3_ are the average hazard ratios in weeks 1–2, weeks 3–4, and weeks 5–10, respectively, under the piecewise constant approximation.

The persons who seek updated vaccines may be intrinsically at higher or lower risk of COVID-19 than the persons who do not seek updated vaccines due to differences in underlying medical conditions, behaviours (e.g., wearing masks, avoiding close contacts), and other personal characteristics (e.g., age, occupation). To remove this confounding bias, we consider a new measure of vaccine effectiveness by comparing HR_3_ to HR_1_

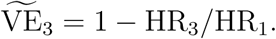

As explained below, this quantity provides a lower bound for the vaccine effectiveness in weeks 5–10 for the population of vaccine seekers, namely the proportionate reduction in the risk of disease in weeks 5–10 when this population is vaccinated compared to when it is not vaccinated.

Let *λ*_1*,V*_ and *λ*_3*,V*_ denote the hazard rates in weeks 1–2 and weeks 5–10, respectively, for vaccinated individuals, and let *λ*_1,*V̄*_ and *λ*_3,*V̄*_ denote the hazard rates in weeks 1–2 and weeks 5–10, respectively, for unvaccinated individuals. Then

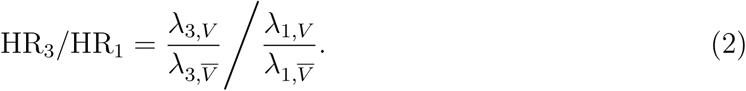

If the level of community transition is constant over time, then *λ*_3,*V̄*_ = *λ*_1,*V̄*_, such that

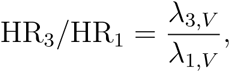

which compares the hazard rates between weeks 5–10 and weeks 1–2. Thus, 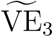 is a lower bound for the effectiveness of updated vaccines for the population of vaccine seekers because the hazard rate would be higher than *λ*_1*,V*_ if this population were not vaccinated.

Let 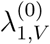 and 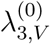 denote the hazard rates that vaccine recipients would have in weeks 1–2 and weeks 5–10, respectively, had they not been vaccinated. Then equation (2) can be written as

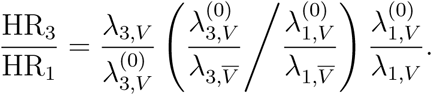

It is reasonable to assume that 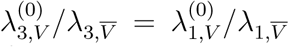, because both ratios measure the relative intrisic risk of COVID-19 for vaccine seekers versus non-seekers due to differences in personal characteristics. Under this assumption,

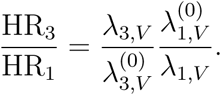

Because the risk of disease for vaccine recipients would be higher if they were unvaccinated, we have 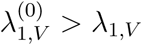, which implies that

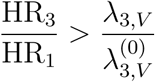

or

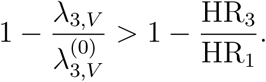

That is, the vaccine effectiveness in weeks 5–10 for the population of vaccine seekers is greater than 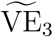.

---

**Figure S1.**
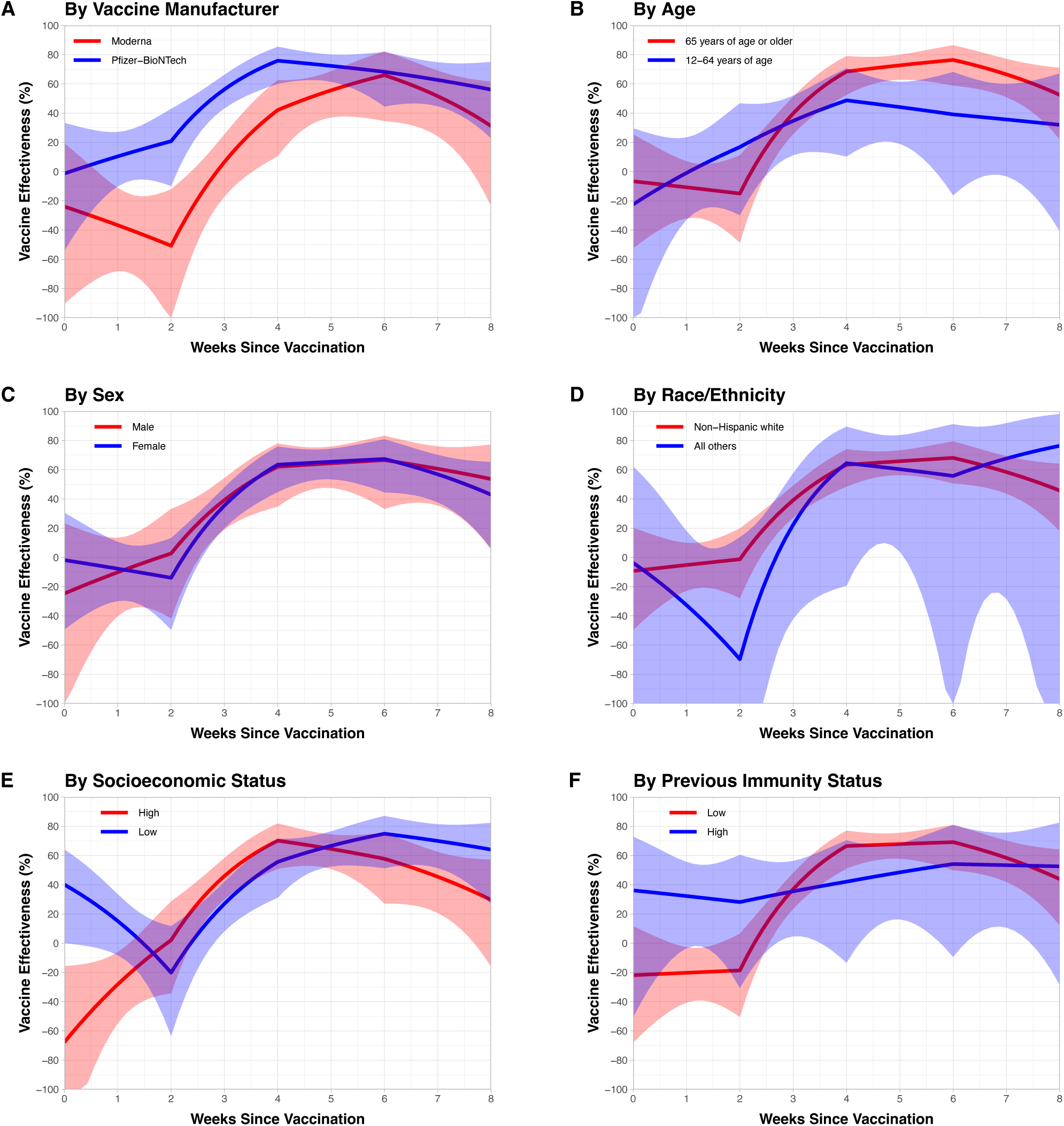
Effectiveness of XBB.1.5 Vaccines in Reducing the Risk of SARS-CoV-2 Infection as a Function of Time Since Vaccination, by Manufacturer and by Demographic and Immunity Subgroups. Estimates are shown by solid curves, and 95% confidence intervals are indicated by shaded bands.

**Table S1.** Estimates (95% CI) for the Effectiveness of XBB.1.5 Vaccines in Reducing the Risk of SARS-CoV-2 Infection, as a Function of Time Since Vaccination, for All Persons.

| **Weeks** | **All Persons** |
| --- | --- |
| 0 | -9.6% (-48.1, 18.8) |
| 1 | -8.6% (-25.8, 6.3) |
| 2 | -7.5% (-34.1, 13.8) |
| 3 | 36.9% (25.1, 46.9) |
| 4 | 63.0% (48.6, 73.4) |
| 5 | 65.1% (55.8, 72.5) |
| 6 | 67.1% (49.9, 78.4) |
| 7 | 58.3% (44.0, 68.9) |
| 8 | 47.1% (20.4, 64.8) |

**Table S2.** Estimates (95% CI) for the Effectiveness of XBB.1.5 Vaccines in Reducing the Risk of SARS-CoV-2 Infection, as a Function of Time Since Vaccination, by Vaccine Manufacturer and Age.

| Weeks | By Vaccine Manufacturer |  | Age |  |
| --- | --- | --- | --- | --- |
|  | Moderna | Pfizer–BioNTech | ≥65 Years | 12–64 Years |
| 0 | -23.9% (-90.4, 19.3) | -1.3% (-54.1, 33.4) | -6.6% (-52.5, 25.4) | -22.4% (-113.1, 29.7) |
| 1 | -36.7% (-68.1, -11.1) | 10.4% (-10.1, 27.1) | -10.7% (-32.0, 7.1) | -0.9% (-32.9, 23.4) |
| 2 | -50.7% (-102.7, -12.1) | 20.8% (-9.9, 42.9) | -15.0% (-48.7, 11.0) | 16.8% (-29.8, 46.7) |
| 3 | 6.6% (-17.0, 25.5) | 56.3% (43.3, 66.3) | 39.8% (25.7, 51.2) | 34.7% (11.3, 51.9) |
| 4 | 42.1% (10.6, 62.6) | 75.9% (60.1, 85.4) | 68.5% (52.5, 79.0) | 48.7% (10.3, 70.7) |
| 5 | 55.6% (37.1, 68.7) | 72.3% (61.5, 80.1) | 72.7% (62.9, 80.0) | 44.1% (19.0, 61.5) |
| 6 | 66.0% (34.6, 82.3) | 68.3% (44.6, 81.8) | 76.4% (58.9, 86.5) | 39.1% (-16.2, 68.1) |
| 7 | 51.6% (24.3, 69.1) | 62.7% (44.8, 74.8) | 66.5% (51.0, 77.1) | 35.7% (-3.2, 59.9) |
| 8 | 31.2% (-23.5, 61.7) | 56.1% (22.8, 75.1) | 52.5% (22.1, 71.0) | 32.0% (-40.9, 67.2) |

**Table S3.** Estimates (95% CI) for the Effectiveness of XBB.1.5 Vaccines in Reducing the Risk of SARS-CoV-2 Infection, as a Function of Time Since Vaccination, by Sex and Race/Ethnicity.

| Weeks | Sex |  | Race/Ethnicity |  |
| --- | --- | --- | --- | --- |
|  | Male | Female | Non-Hispanic White | All Others |
| 0 | -24.7% (-102.8, 23.3) | -1.8% (-49.3, 30.5) | -9.3% (-49.7, 20.3) | -3.7% (-184.4, 62.2) |
| 1 | -10.1% (-40.2, 13.4) | -7.7% (-29.8, 10.6) | -5.2% (-22.8, 9.9) | -32.6% (-114.5, 18.0) |
| 2 | 2.7% (-41.7, 33.2) | -13.9% (-49.6, 13.2) | -1.3% (-27.9, 19.8) | -69.6% (-233.8, 13.8) |
| 3 | 39.3% (19.2, 54.3) | 35.5% (20.0, 47.9) | 39.1% (27.2, 49.1) | 22.4% (-42.1, 57.6) |
| 4 | 62.1% (34.8, 77.9) | 63.5% (44.7, 75.9) | 63.4% (48.5, 74.0) | 64.5% (-19.4, 89.4) |
| 5 | 64.4% (47.5, 75.8) | 65.5% (53.5, 74.4) | 65.8% (56.3, 73.3) | 60.4% (4.0, 83.6) |
| 6 | 66.6% (33.1, 83.3) | 67.4% (44.5, 80.8) | 68.1% (50.7, 79.4) | 55.8% (-120.9, 91.1) |
| 7 | 60.6% (35.5, 76.0) | 56.8% (37.7, 70.1) | 58.4% (43.6, 69.3) | 67.7% (-28.3, 91.9) |
| 8 | 53.6% (5.5, 77.3) | 42.9% (6.0, 65.3) | 45.7% (17.9, 64.1) | 76.4% (-188.4, 98.1) |

**Table S4.** Estimates (95% CI) for the Effectiveness of XBB.1.5 Vaccines in Reducing the Risk of SARS-CoV-2 Infection, as a Function of Time Since Vaccination, by Socioeconomic and Previous Immunity Status.

| Weeks | Socioeconomic Status |  | Previous Immunity Status |  |
| --- | --- | --- | --- | --- |
|  | High | Low | High | Low |
| 0 | -67.7% (-143.2, -15.6) | 40.1% (0.0, 64.1) | 36.2% (-50.5, 72.9) | -21.8% (-68.0, 11.6) |
| 1 | -28.1% (-54.8, -6.0) | 15.2% (-8.2, 33.5) | 32.3% (-3.4, 55.6) | -20.2% (-40.6, -2.8) |
| 2 | 2.1% (-34.3, 28.7) | -20.1% (-63.3, 11.7) | 28.1% (-30.6, 60.5) | -18.6% (-50.3, 6.4) |
| 3 | 46.0% (30.0, 58.3) | 27.0% (8.2, 42.0) | 35.5% (4.7, 56.3) | 37.0% (23.5, 48.1) |
| 4 | 70.2% (50.9, 81.9) | 55.7% (31.2, 71.4) | 42.1% (-13.3, 70.4) | 66.5% (51.1, 77.1) |
| 5 | 64.5% (50.8, 74.4) | 66.7% (52.5, 76.6) | 48.5% (16.3, 68.3) | 67.9% (57.8, 75.5) |
| 6 | 57.8% (27.2, 75.5) | 74.9% (51.3, 87.1) | 54.2% (-9.4, 80.8) | 69.2% (50.0, 81.0) |
| 7 | 45.5% (20.7, 62.5) | 70.0% (51.5, 81.4) | 53.4% (10.7, 75.7) | 58.4% (41.9, 70.2) |
| 8 | 29.6% (-16.0, 57.3) | 64.1% (26.8, 82.4) | 52.6% (-28.7, 82.5) | 43.9% (12.2, 64.1) |

